# A simple model to fit the time evolution of the daily death rate of Covid-19 in European Union countries

**DOI:** 10.1101/2020.05.06.20093062

**Authors:** Tristan Beau, Julien Browaeys, Olivier Dadoun

## Abstract

We suggest a minimal model to describe the evolution of daily deaths due to Covid-19 in the European Union (EU) and UK, without any epidemiological hypothesis. Assuming current lockdown conditions were to remain in place, as of May 3rd 2020, this *ad hoc* fitting model is forecasting a total of 204 586 deaths in the EU. We could currently be at 2/3 of the total casualty count, and reach an overall death rate of one over 2 500 people.

## I. — Introduction

The current Covid-19 pandemia is a major worldwide concern since its outbreak in December 2019[1]. Although all countries are publishing their statistics in term of deaths, confirmed or recovered cases, the epidemiological modeling remains complex. In this study, we propose an modelization of data without any epidemiological assumptions.

The realization of a major epidemic crisis happening in the European Union (EU) came in March 2020, and most EU countries consequently instituted some form of lockdown in a matter of days. Since then, at least in Europe, the epidemics has receded enough so that these countries have planned to lift the lockdown, at least partially, in the second week of May.

We wish to contribute to the evaluation of the incidence of such a decision based on current data. We fit an effective physical model to public Covid-19 deaths data in all 28 EU countries (including UK) in order to forecast the evolution of the epidemic if lockdown was maintained. In that case, the total number of deaths is estimated.

## II. — Data and method

The John Hopkins University database[5] provides a daily report of deaths, confirmed and recovered Covid-19 patients all over the world, since January 22nd, 2020; this date is considered to be our time reference *t* = 0. Because of large variations in testing policies in different countries, in this analysis we limit ourselves to the study of daily deaths as a function of time. We are aware that this remains an imperfect proxy of the epidemic as counting policies can also vary from one country to the other.

In order to be able to compare countries, we normalize data by country population. This information is taken from the latest *United Nations Population Division* estimates [4]. Databases extraction of data and their initial analysis is ensured by the open source software called CoCoA[2, 3].

## III. — Analysis and extrapolation

The effective modeling of data is done by a standard least-square fit of a Crystal Ball function[6].

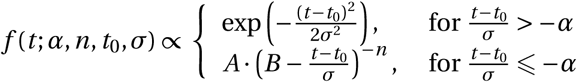

The Crystal Ball function consists of a smoothly joined Gaussian part and a power-law tail at a certain cut-off. It has 5 parameters: *N* is the full integral of the function over the time, *n* the power law parameter, *α* a normalized cutoff where the power-law replaces the Gaussian part, *t*_0_ the position of the peak, *σ* the time scale. We are only reporting the useful parameters *N* and *t*_0_, which respectively describe the total number of death (assuming the daily death rate to follow this function) and the time at which the peak of the epidemic occurred.

Commonly used in high-energy physics, this function is named after a neutral particle detector initially used at the Stanford Linear Accelerator Center. The reason for choosing this function is two-fold. First we needed to describe properly the asymmetrical bell shape curve of the data (see figures below). Second we wanted a minimum number of adjustable parameters to avoid over-fitting.

For every EU countries (and UK), we represent daily deaths as a function of time since *t*=0, together with the fitted Crystal Ball function. It is represented over 200 days to better visualize our forecast. The best fit parameters *N* and *t*_0_ are given on each figure.

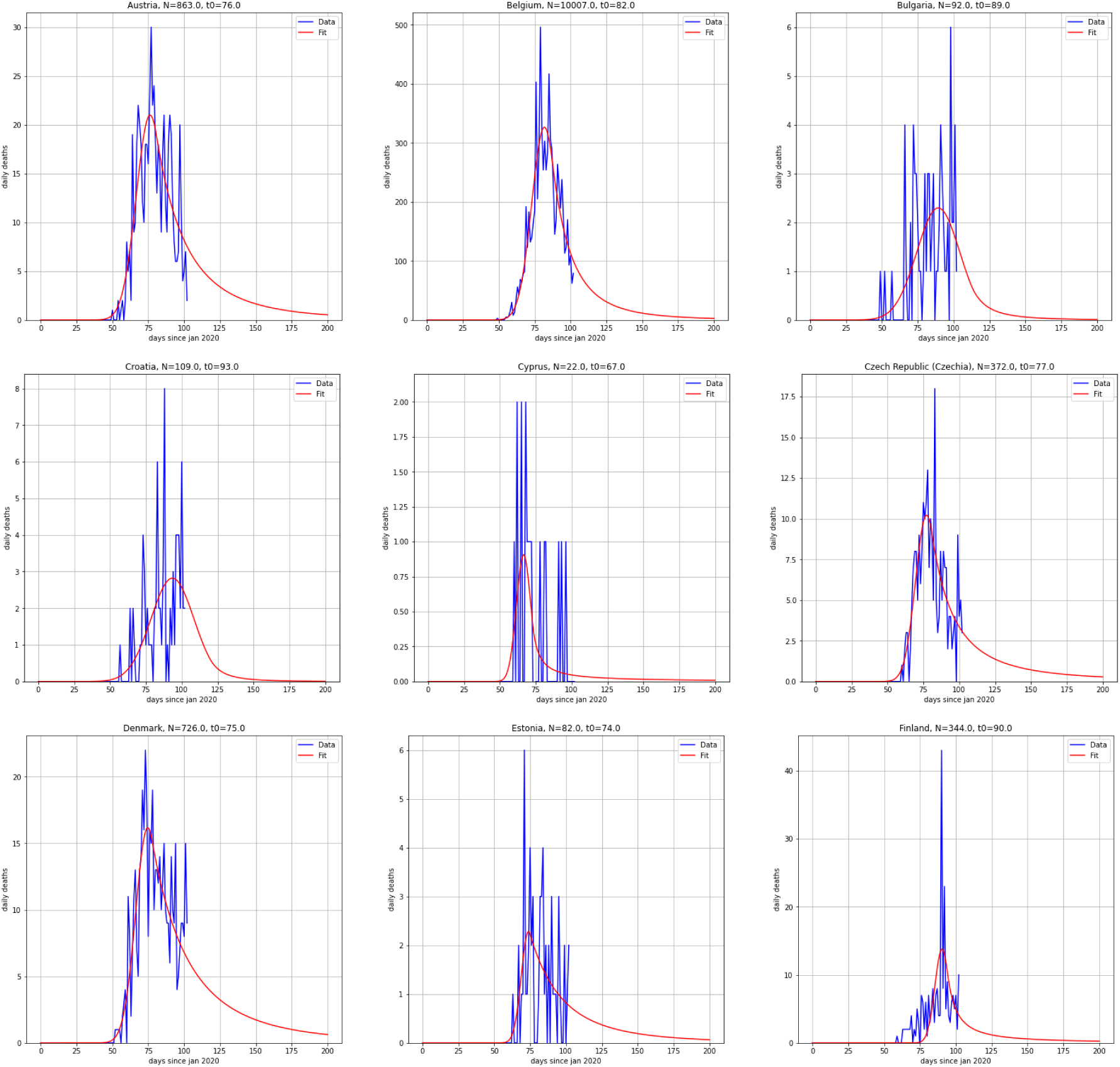

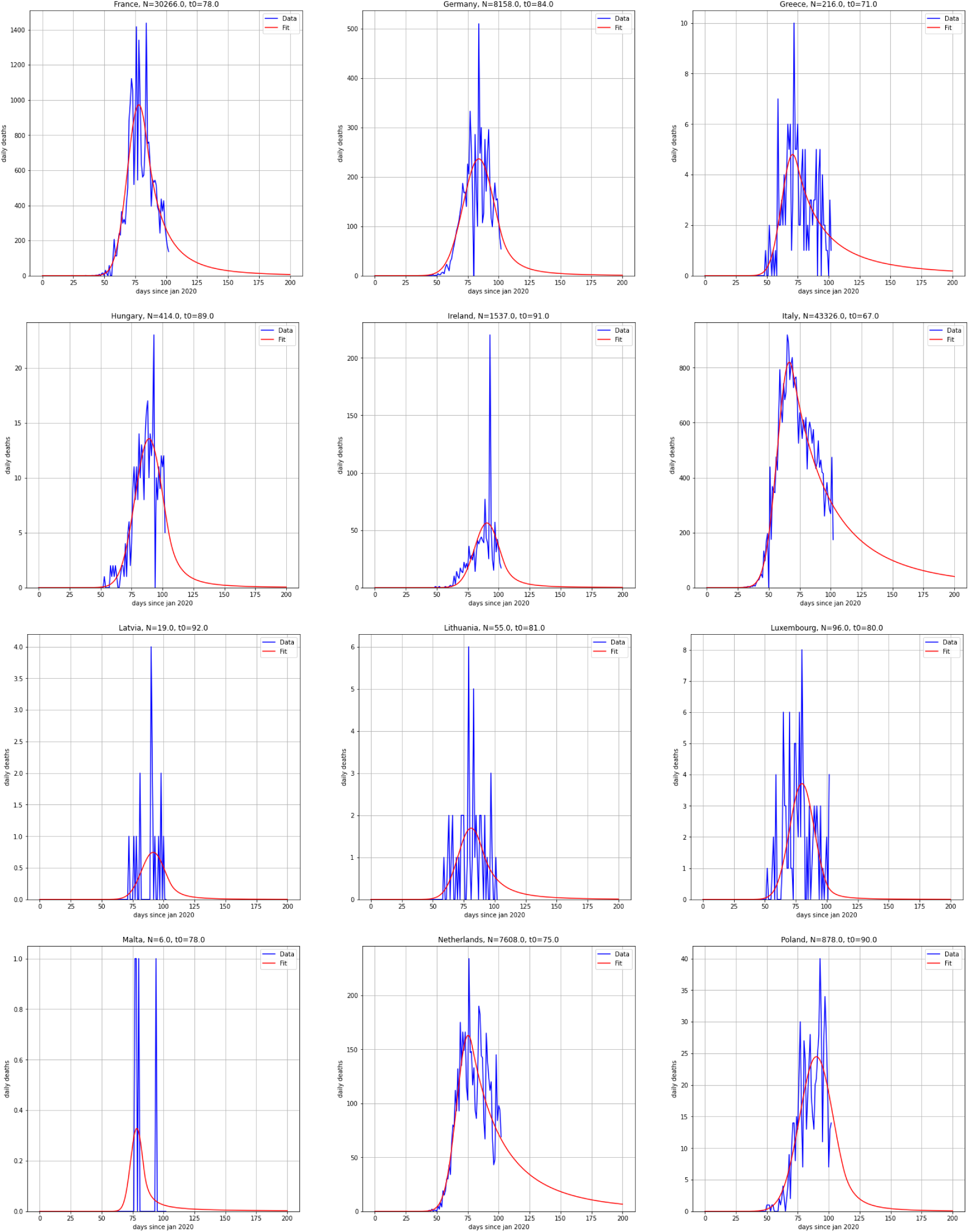

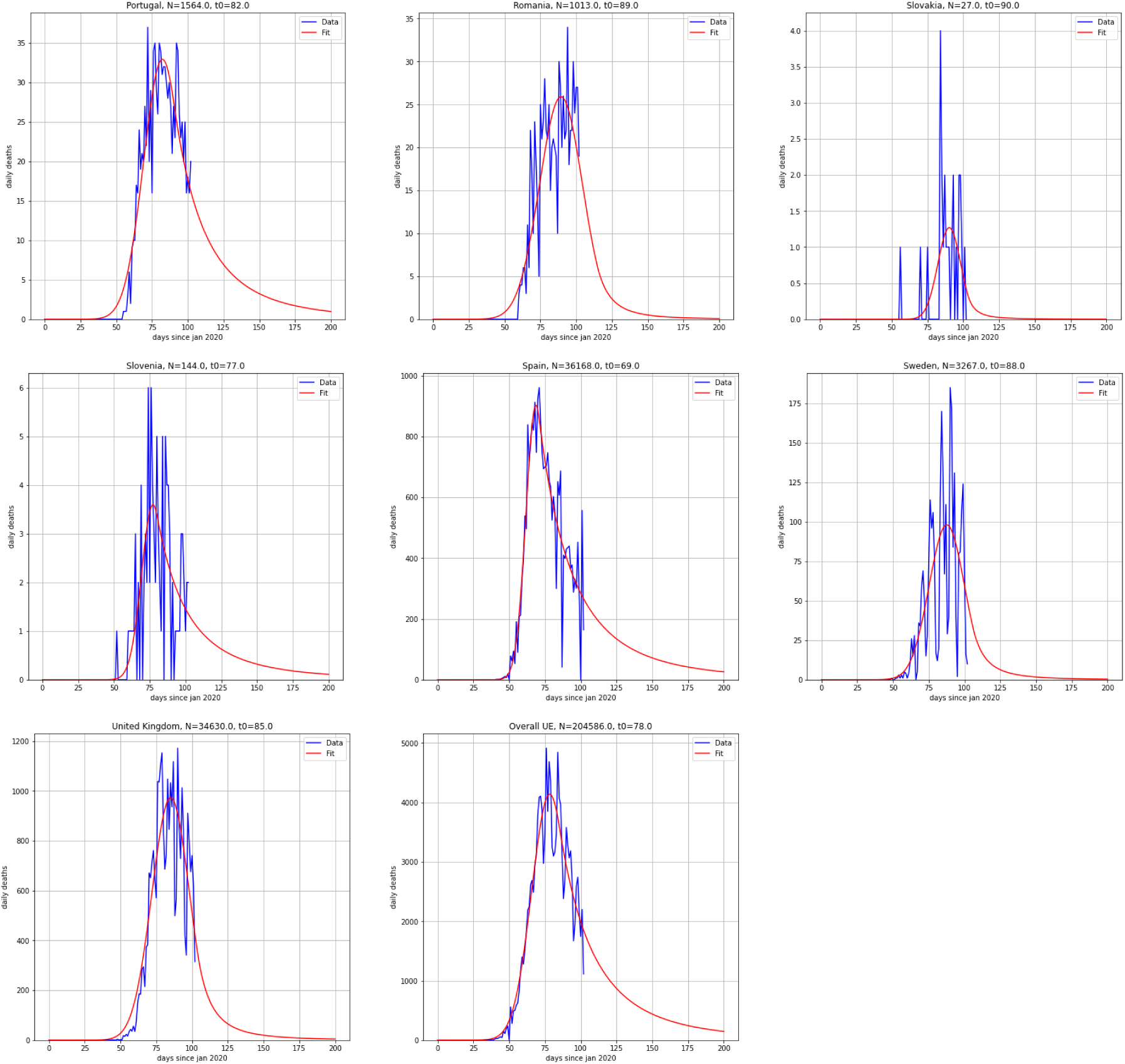

The same procedure has also been applied to European Union (including UK) as a whole.

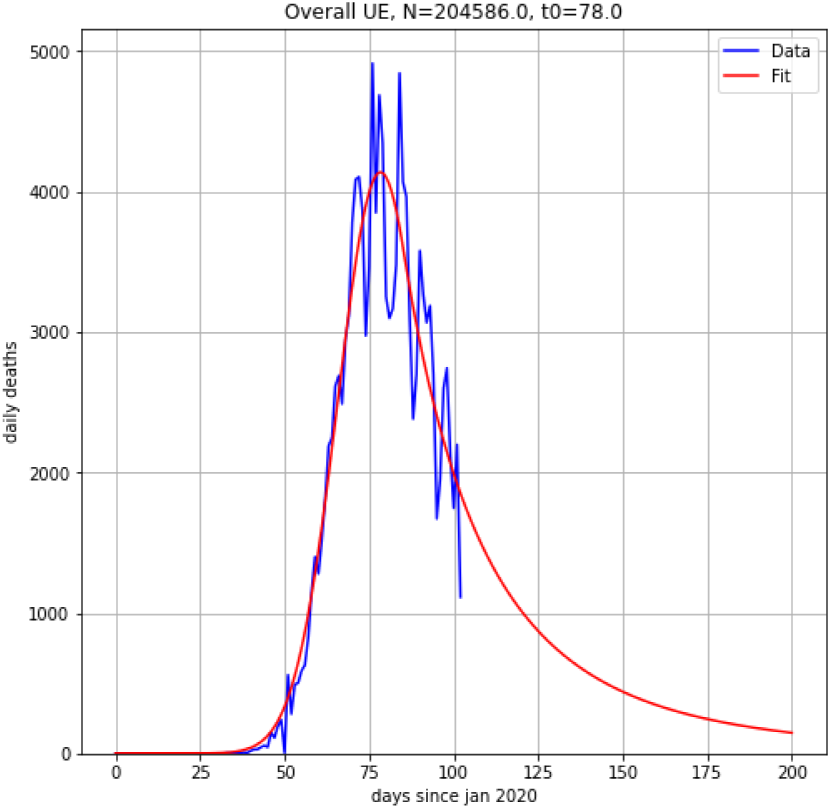

## IV. — Discussion

Our modeling yields an estimation of the expected overall number of deaths for each country. We provide these numbers in the table below, and compare them with the total number deaths reported until May 3rd, 2020, to infer the progress of the epidemic at this date. We normalize the figures by population to facilitate comparison between countries.

| Country | Population | Deaths<br>(May<br>3rd) | Total<br>forecast<br>$N$ | Peak<br>date $t_0$ | Deaths<br>(May<br>3rd) per<br>100k pop | Total<br>forecast<br>per 100k<br>pop | Extent<br>(May<br>3rd) in % |
| --- | --- | --- | --- | --- | --- | --- | --- |
| UE + UK | 513 136 525 | 137 796 | 204 586 | 78 | 26.9 | 39.9 | 67.4 |
| Germany | 83 783 942 | 6 993 | 8 158 | 84 | 8.3 | 9.7 | 85.7 |
| United Kingdom | 67 886 011 | 28 809 | 34 630 | 85 | 42.4 | 51.0 | 83.2 |
| France | 65 273 511 | 25 204 | 30 266 | 78 | 38.6 | 46.4 | 83.3 |
| Italy | 60 461 826 | 29 079 | 43 326 | 67 | 48.1 | 71.7 | 67.1 |
| Spain | 46 754 778 | 25 428 | 36 168 | 69 | 54.4 | 77.4 | 70.3 |
| Poland | 37 846 611 | 698 | 878 | 90 | 1.8 | 2.3 | 79.5 |
| Romania | 19 237 691 | 818 | 1 013 | 89 | 4.3 | 5.3 | 80.8 |
| Netherlands | 17 134 872 | 5 098 | 7 608 | 75 | 29.8 | 44.4 | 67.0 |
| Belgium | 11 589 623 | 7 924 | 10 007 | 82 | 68.4 | 86.3 | 79.2 |
| Czech Rep. | 10 708 981 | 252 | 372 | 77 | 2.4 | 3.5 | 67.7 |
| Greece | 10 423 054 | 146 | 216 | 71 | 1.4 | 2.1 | 67.6 |
| Portugal | 10 196 709 | 1 063 | 1 564 | 82 | 10.4 | 15.3 | 68.0 |
| Sweden | 10 099 265 | 2 769 | 3 267 | 88 | 27.4 | 32.3 | 84.8 |
| Hungary | 9 660 351 | 351 | 414 | 89 | 3.6 | 4.3 | 84.8 |
| Austria | 9 006 398 | 600 | 863 | 76 | 6.7 | 9.6 | 69.5 |
| Bulgaria | 6 948 445 | 78 | 92 | 89 | 1.1 | 1.3 | 84.8 |
| Denmark | 5 792 202 | 493 | 726 | 75 | 8.5 | 12.5 | 67.9 |
| Finland | 5 540 720 | 240 | 344 | 90 | 4.3 | 6.2 | 69.8 |
| Slovakia | 5 459 642 | 25 | 27 | 90 | 0.5 | 0.5 | 92.6 |
| Ireland | 4 937 786 | 1 319 | 1 537 | 91 | 26.7 | 31.1 | 85.8 |
| Croatia | 4 105 267 | 80 | 109 | 93 | 1.9 | 2.7 | 73.4 |
| Lithuania | 2 722 289 | 46 | 55 | 81 | 1.7 | 2.0 | 83.6 |
| Slovenia | 2 078 938 | 97 | 144 | 77 | 4.7 | 6.9 | 67.4 |
| Latvia | 1 886 198 | 16 | 19 | 92 | 0.8 | 1.0 | 84.2 |
| Estonia | 1 326 535 | 55 | 82 | 74 | 4.1 | 6.2 | 67.1 |
| Cyprus | 1 207 359 | 15 | 22 | 67 | 1.2 | 1.8 | 68.2 |
| Luxembourg | 625 978 | 96 | 96 | 80 | 15.3 | 15.3 | 100.0 |
| Malta | 441 543 | 4 | 6 | 78 | 0.9 | 1.4 | 66.7 |

With merely five free parameters, our model fits reasonably well all data for the 28 EU countries (and UK). Fitting seems all the more accurate that the peak can be easily identified, and that the total number of daily death is high.

Some countries plan lifting the lockdown in early May 2020, such as Germany (May 4th), or France (May 11th). One can assume that without lifting the lockdown, the evolution of the daily rate of death could have followed the trend we have exhibited.

On May 3rd, EU countries (and UK) have recorded a total of 136 391 deaths due to Covid-19. Our effective model is projecting 204 586 deaths if all conditions remain as current. In this hypothesis, 2/3 of the total number of Covid-19 victims have already perished. It will be interesting to compare these figures with the total number of death at the end of the epidemic.

We should stress that our proposal is prospective and not predictive, in the sense that it does *not* rely on epidemiological modeling. We merely describe data through *ad hoc* fitting of mathematical functions, a methodology similar to what is used in economic forecasts.

## V. — Conclusion

We provide an minimal model, within the CoCoA framework, that reasonably fits the daily rate of death as a function of time with only five free parameters. Assuming lockdown is not lifted, we estimate that at the beginning of May 2020 about two third of deaths have already occurred in Europe before the end of the epidemic. This implies a final total death rate of 1 over 2 500 people. In the future, lockdown lifting policies could be evaluated with regard to our projection.

## Data Availability

The data that support the findings of this study are available in CSSEGISandData/COVID-19 by the Center for Systems Science and Engineering (CSSE) at Johns Hopkins University, at [https://github.com/CSSEGISandData/COVID-19]. And also in Population by Country (2020) - Worldometer, at [https://www.worldometers.info/world-population/population-by-country/]. These data were derived from the following resources available in the public domain: [https://github.com/tjbtjbtjb/CoCoA].

